# Retinoblastoma Detection via Image Processing and Interpretable Artificial Intelligence Techniques

**DOI:** 10.1101/2023.05.02.23289419

**Authors:** Surya Duraivenkatesh, Aditya Narayan, Vishak Srikanth, Adamou Fode Made

**Affiliations:** Stanford Online High School, San Ramon, US; Dougherty Valley High School, San Ramon, US; Stanford Online High School, Redwood City, US; Stanford Online High School, Stanford University, Redwood City, US

**Keywords:** Retinoblastoma, LMICs, VGG16, VGG19, Xception, Inception v3, Xception, DSC, IoU, SHAP, LIME

## Abstract

Retinoblastoma (RB) is a treatable ocular melanoma that is diagnosed early and subsequently cured in the United States but has a poor prognosis in low- and middle-income countries (LMICs). This study outlines an approach to aid health-care professionals in identifying RB in LMICs. Transfer learning methods were utilized for detection from fundus imaging. One hundred and forty RB+ and 140 RB-images were acquired from a previous deep-learning study. Next, five models were tested: VGG16, VGG19, Xception, Inception v3, and ResNet50, which were trained on the two-hundred-and-eighty image dataset. To evaluate these models, the Dice Similarity Coefficient (DSC) and Intersection-over-Union (IoU) were used. Explainable AI techniques such as SHAP and LIME were implemented into the best-performing models to increase the transparency of their decision-making frameworks, which is critical for the use of AI in medicine. We present that VGG16 is the best at identifying RB, though the other models achieved great levels of prediction. Transfer learning methods were effective at identifying RB, and explainable AI increased viability in clinical settings.

## I. Exordium

Retinoblastoma (RB) is a treatable ocular melanoma and is considered an orphan disease (impacts less than 200,000 people nationwide) caused by a mutation in both RB1 genes within a cell (The American Cancer Society Medical and Editorial content team, 2018). RB typically occurs under 2 years of age and is diagnosed early and subsequently cured in the United States but disproportionately impacts low- and middle-income countries (LMICs). In a study where 4064 patients with RB participated, 642 (15.8%) were from high-income countries, 1151 (28.3%) were from upper-middle-income countries, 1791 (44.1%) from lower-middle-income countries, and 480 (11.8%) from low-income countries (Fabian et al., 2022). When going from high- to low-income countries, the percentage of mortality significantly increases. This difference in death percentages can be most directly attributed to the lack of specialized treatment centers and equipment needed to identify RB early on, leading to the further spread of RB outside of the eye.

### A. Engineering Goals

Current machine learning work on the topic of Retinoblastoma has reasonable accuracy. However, in clinical settings, accuracy is never sufficient until perfect. This project wishes to advance RB diagnostic accuracy by utilizing transfer learning techniques. Explainable AI (XAI) is also an unexplored sector of RB research; this project also wishes to use XAI to increase the informational applications of the models to help doctors identify unique tumor features, alongside making the CNN model’s process as transparent as possible to maximize its applications in clinical settings.

**TABLE I.** RB mortality rate by income class; info from (Fabian et al.)

| Income Class | Numerical Data |  |  |
| --- | --- | --- | --- |
|  | Number of People | # of Deaths | Percent of Death |
| High | 642 | 5 | 0.8% |
| Upper-Middle | 1151 | 92 | 8% |
| Lower-Middle | 1791 | 276 | 15.4% |
| Low | 480 | 146 | 30.4% |
| Totals | 4064 | 519 | X |

## II. Transfer Learning

Transfer learning is a powerful tool that utilizes a pre-trained model on a new problem; this approach is accommodating when obtaining a large dataset is not possible. In this analysis, the pre-trained models VGG16, VGG19, ResNet50, Inception v3, and Xception are compared. These models were originally trained with the ImageNet dataset but were later trained on our 280-image dataset. A biomedical segmentation CNN such as U-net was considered, but there exists a lack of transparency; U-net provides the diagnosis and area of impact but does not provide sufficient detail. With U-net, it is difficult to explore novel diagnostic tumor features due to the generality of the segmentation model, but with XAI techniques, it is possible to find specific, and often minute details of the fundus image that inform the model of a benign/malignant diagnosis.

### A. Data Analysis

Fundus imaging data that pictures RB is scarce online. However, utilizing a previous deep learning study, 140 RB-positive fundus images were obtained (Jeba 2023). To find 140 RB-negative fundus images, research papers were scraped to find healthy benign images. Although this may be a source of error due to differences in how the fundus images were taken, this was the best dataset that could be acquired. Since all models were run with the same dataset, we believe that this did not cause drastic change in which architecture performed the best. Data-manipulating such as data augmentation was considered, but was ultimately decided against as medical imaging is more high-stakes than other image classification tasks. If the data augmentation causes any slight resolution issues, it can dramatically impact the accuracy of predictions. The RB-positive images were captured using a Retcam pediatric camera with patients between the ages 12 to 20 years. The RB-negative images may not have all been taken in the same manner. All images were resized to the proper input size during image and label preprocessing. Although the tumor can be easily distinguished when looking at figures 2 and 3, in other cases, it isn’t as easy.

**Fig. 1.**
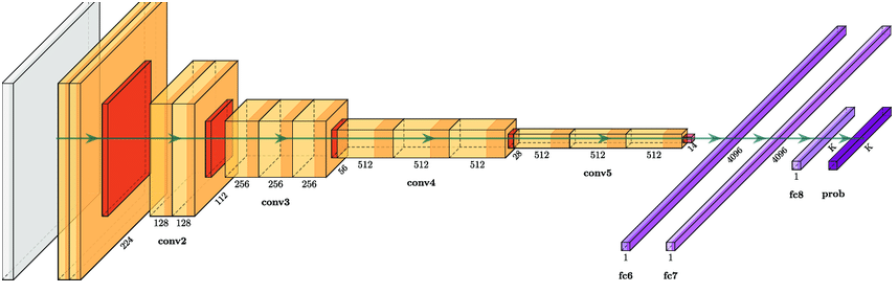
VGG16 architecture; info taken from (Blauch et al., 2021)

**Fig. 2.**
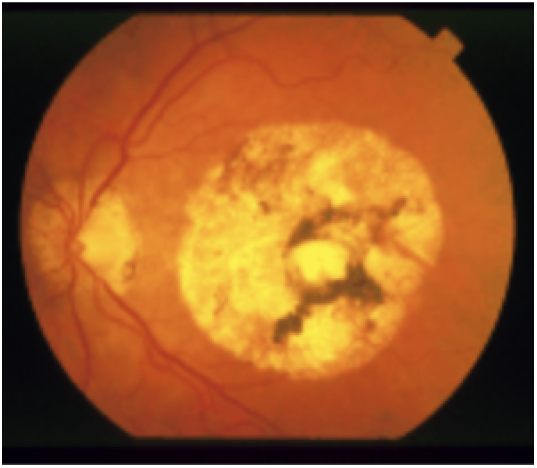
RB-positive image from the dataset

**Fig. 3.**
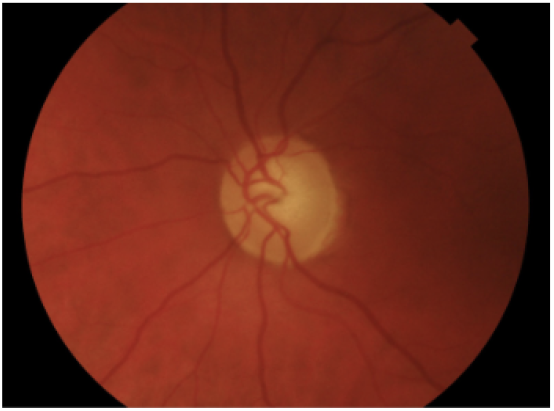
RB-negative image from the dataset

### B. Preliminary Testing

Scikit-learn’s tool Grid Search CV was considered to find the best hyperparameters but was not since it is less feasible with neural networks like the ones used. Though accuracy was not the final metric used, it was utilized for preliminary testing. The concern of overfitting arose since high accuracy rates were obtained. Since all five models returned a similar testing/validation accuracy/loss, it was deduced that overfitting was not a major concern. Early stopping was still implemented to account for overfitting, and the test dataset was used to confirm the absence of overfitting. Since the test received similar results as both training & validation, we concluded that overfitting was not a major issue. For all of the models, the validation split was 0.4 and the patience for early stopping was 5, with validation accuracy being monitored. Epochs were typically 50 (though the model stopped short due to early stopping), with ResNet50 having 4 epochs due to overfitting occurring particularly early. Pre-trained image processing models often take a long time to run, so the epoch count was lowered in preliminary testing. It was later increased when further testing was done.

**Fig. 4.**
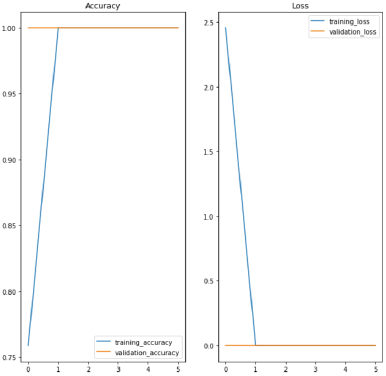
Results with preliminary VGG16 testing

**Fig. 5.**
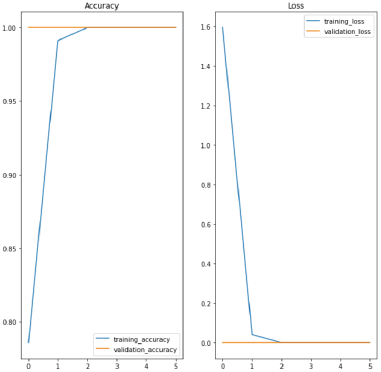
Results with preliminary VGG19 testing

**Fig. 6.**
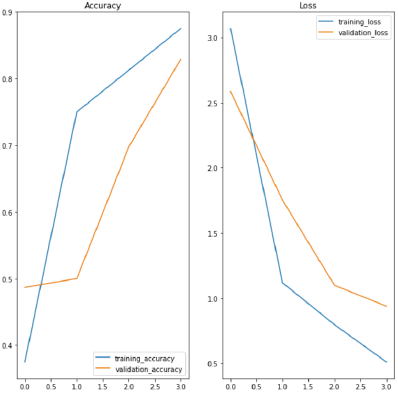
Results with preliminary ResNet50 testing

### C. Metrics

For efficient transfer learning model comparisons, a good way to compare the models is needed. Some metrics that were considered were accuracy, specificity, and ROC AUC. Accuracy isn’t the best indicator, as in a sample of 100 patients, marking all patients as having benign growths if there were 10 patients with cancer would yield a misleading 0.9 accuracy rate. After careful review, it was decided to use Dice Similarity Metric (DSC, also known as F-score) and Intersection-over-Union (IoU), which are considered state-of-the-art metrics for medical image classification. DSC is calculated from a harmonic mean of the precision and recall of a prediction. IoU, also known as the Jaccard index, is another metric that is recently discovered to have very good potential in evaluating a model for its efficacy, with respect to the detection of diseases. IoU typically penalizes under and over-segmentation more than DSC. As the name suggests, IoU is calculated by taking the intersection, and union, and dividing them. In both DSC and IoU, it is noticed that a bigger weightage is placed on true positives; the more true positives and fewer false negatives that a model gives, the higher its Jaccard index and resultant plausibility in clinical applications is.

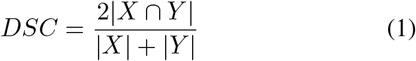

**Fig. 7.**
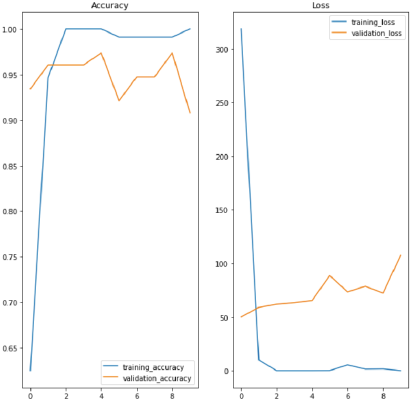
Results with preliminary Inception v3 testing

**Fig. 8.**
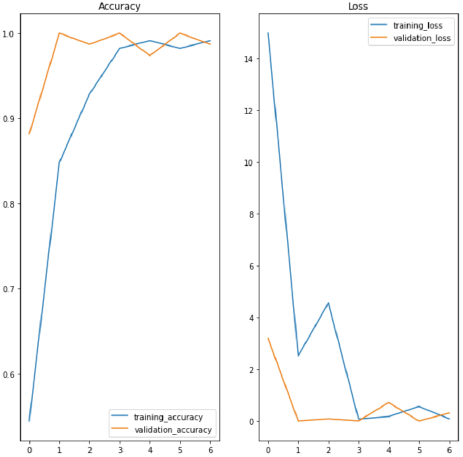
Results with preliminary VGG16 testing

#### (1) DSC formula; info from (“Sørensen–Dice coefficient”, 2023)

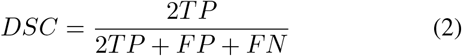

#### (2) DSC formula for binary classification

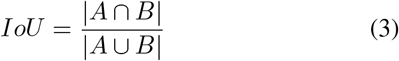

#### (3) IoU formula; info from (“Jaccard index”, 2023)

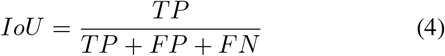

#### (4) IoU formula for binary classification

Utilizing PyTorch’s Jaccard function, the Jaccard index/IoU of every model was taken. The average type was set to micro (calculates metrics globally, across all samples and classes) and converted the parameters to meet the parameter specifications needed to input in the y true and y pred. For y pred, a plethora of functions was run in order to receive the values in a format that was rendered usable by the model.

**Fig. 9.**
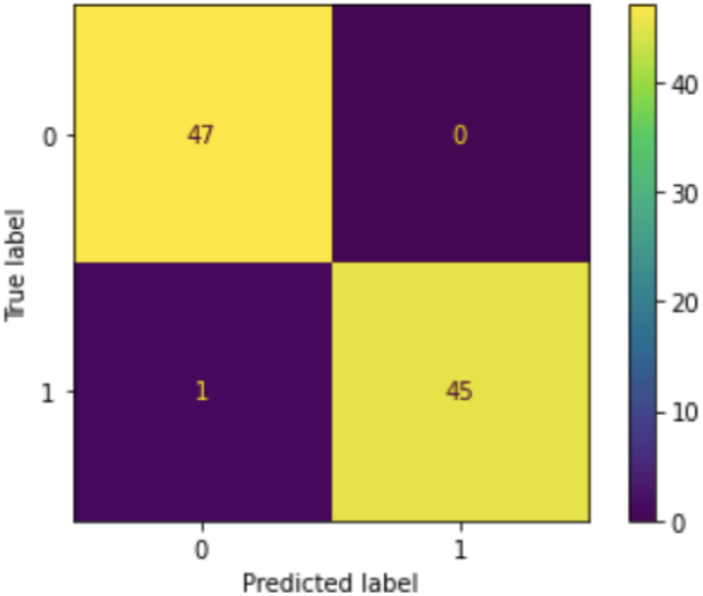
Confusion matrix for VGG16

**Fig. 10.**
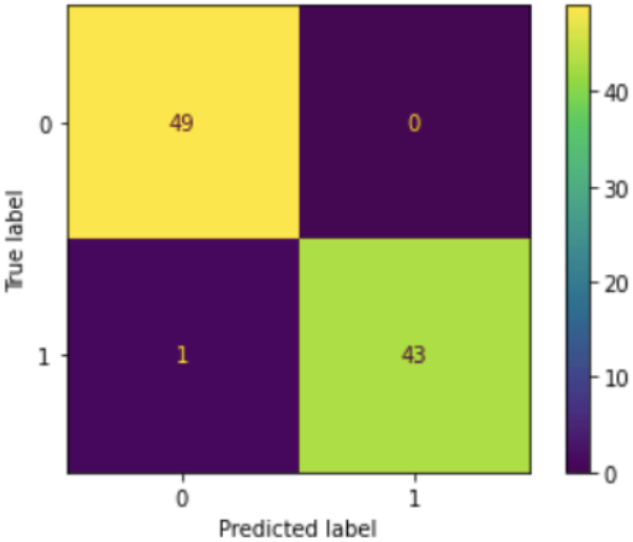
Confusion matrix for VGG19

**TABLE II.**
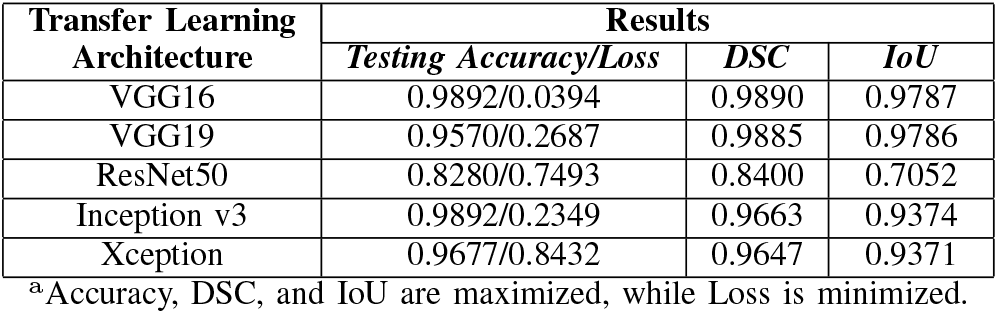
Results with metrics

| Transfer Learning<br>Architecture | Results |  |  |
| --- | --- | --- | --- |
|  | Testing Accuracy/Loss | DSC | IoU |
| VGG16 | 0.9892/0.0394 | 0.9890 | 0.9787 |
| VGG19 | 0.9570/0.2687 | 0.9885 | 0.9786 |
| ResNet50 | 0.8280/0.7493 | 0.8400 | 0.7052 |
| Inception v3 | 0.9892/0.2349 | 0.9663 | 0.9374 |
| Xception | 0.9677/0.8432 | 0.9647 | 0.9371 |

<sup>a</sup>Accuracy, DSC, and IoU are maximized, while Loss is minimized.

With VGG16, VGG19, and ResNet50, the largest concern was overfitting. Often, after just a few epochs, perfect accuracy was achieved, and that provided uncertainty about the validity of these results. Since the validation and training results for accuracy and loss were both similar, clarification was needed about whether or not the models were faced with a situation of overfitting; one of the most common deep learning problems. This issue was mitigated with early stopping, a mechanism that monitors a metric, which in this case was validation accuracy, and stops the training early if it notices a trend of overfitting. In the ResNet model, the model was stopped at 4 epochs, before the early stopping mechanism, since it was noticed that the ResNet50 model progressively declined past 4 epochs. However, in VGG16 and VGG19, the early stopping mechanism was enough to stop the model at a very high accuracy. In addition, to validate the results given by the validation accuracy and loss, a testing dataset was separated at the beginning of running each model and utilized to receive an accuracy and loss value; in all models, these came out similar to the results of the training and validation, giving us reason to believe that there was no overfitting after the measures taken. As VGG16 and VGG19 are state-of-the-art image-processing models, it is reasonable to see high-accuracy results in binary classification tasks such as these. The second concern was the long time to run these models; since there were many layers to get through, often, running these models would take a very long time. This was mitigated by either reducing epochs if the current stage was preliminary testing or the model training was run in the background while other aspects of the project were completed.

**Fig. 11.**
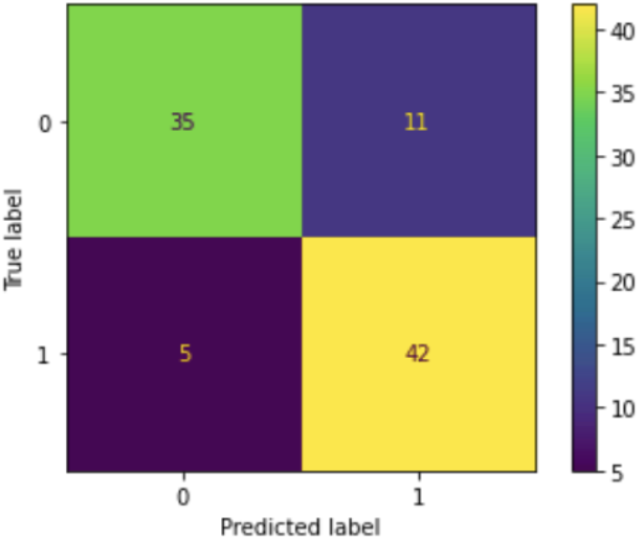
Confusion matrix for ResNet50

**Fig. 12.**
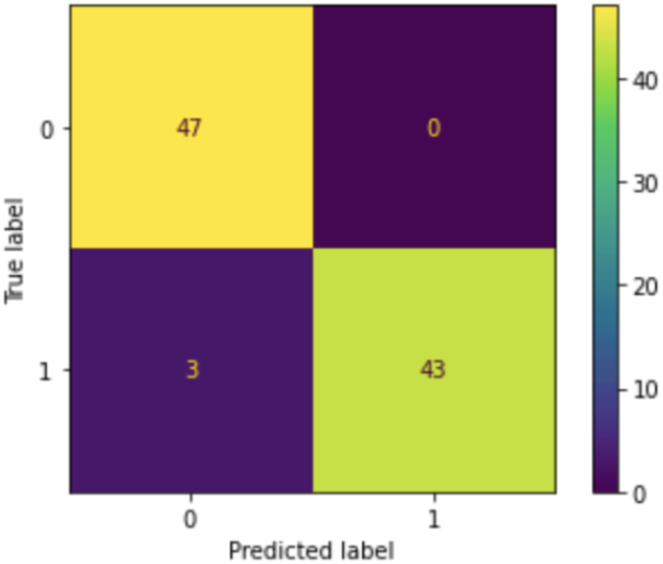
Confusion matrix for Inception v3

**Fig. 13.**
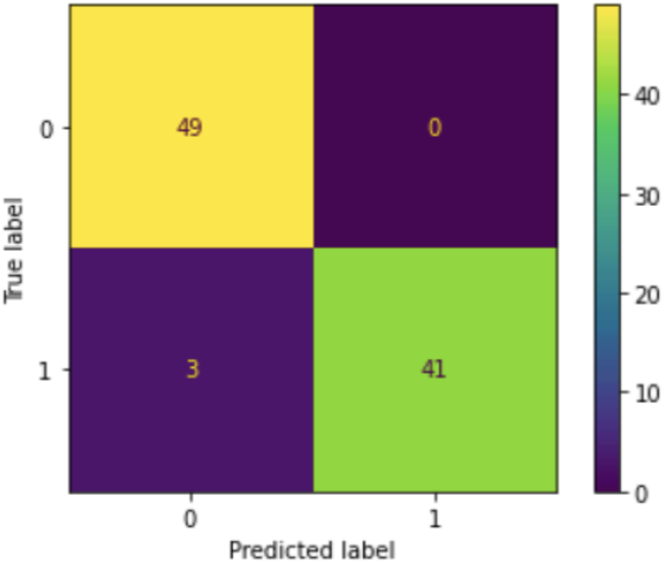
Confusion matrix for Xception

It was noticed that the Inception v3 model had a relatively good Jaccard index and DSC but had an unfathomably high loss amount. It is believed that the lack of a lot of training data caused the model to pick up on noise that wasn’t necessarily significant. It is believed that the Inception v3 model has good feasibility for usage in healthcare if the data input were to increase so that the variety of layers in the model wouldn’t catch noise as easily.

Input size drastically affected the results of models. ResNet50 takes an input size of 224×224, but at first, 64×64 was selected by accident. This drastically affected the results since the benign images were resized to much smaller dimensions. In addition, for ResNet50, this means that the malignant images were scaled down rather than scaled up. These contributed to the initial better results for ResNet50, which are attached. For ResNet50 specifically, although early stopping measures had already been in place, a force stop at 4 epochs occurred due to overfitting happening extremely early on. With this extra measure, decent results were achieved.

**TABLE III.** Initial results for main ResNet50 testing due to input size

| Transfer Learning Architecture | Results |  |  |
| --- | --- | --- | --- |
|  | Testing Accuracy/Loss | DSC | IoU |
| ResNet50 | 0.8817/0.8578 | 0.8866 | 0.7881 |

<sup>a</sup>Accuracy, DSC, and IoU are maximized, while Loss is minimized.

In summary, there were various factors that led up to the results that were received. Most notably overfitting and model complexity, but regardless it is deduced that the VGG16 model is the fittest model for RB detection. VGG19 was a very close runner-up to VGG16. Although the VGG19 model has a possibility to have pushed over the tiny margin it missed with more data, the same could be argued for all the models.

**Fig. 14.**
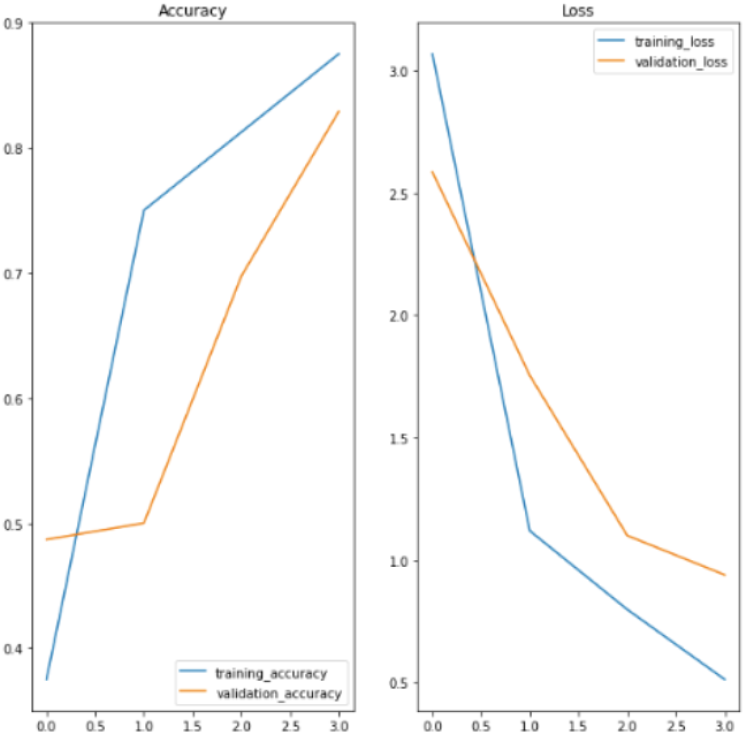
Initial results for preliminary ResNet50 testing due to input size

**Fig. 15.**
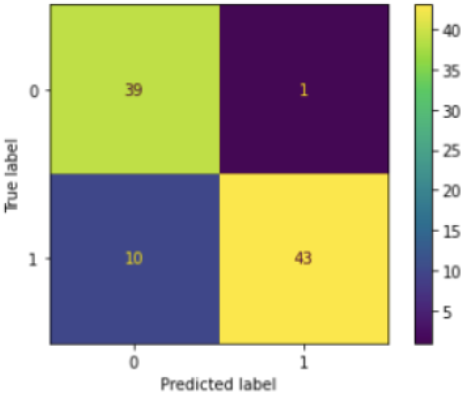
Initial confusion matrix for ResNet50 testing due to input size

## III. Interpretable Artificial Intelligence

This project wishes to maximize the clinical applications of the architecture that was discovered to be most effective in RB detection. However, in medicine, after accuracy is transparency, a significant factor that influences a lot of decisions. Often AI is not trusted due to the black-box setup that is forced upon doctors when they attempt to utilize AI in healthcare applications. One solution to this is the usage of the novel explainable AI techniques of SHapley Additive Explanations (SHAP) and Local Interpretable Model Agnostic Explanations (LIME) come in handy. They help provide insight into which part of the image informed the model of the presence of RB. LIME wants to minimize the locality-aware loss L(f,g,?x) while keeping the model complexity denoted low. SHAP was the result of the VGG16 model during preliminary testing and was used to predict a single malignant image. First, the image was segmented so each pixel didn’t need to be explained. Then, the top prediction was retrieved. After visualizing the explanations, an image with SHAP values on it was obtained. The SHAP value talks about how much each value contributed toward the final outcome. A negative SHAP value for a feature means that the feature contributed more toward the listed diagnosis, and a positive SHAP value for a feature means that the feature contributed more away from the listed diagnosis.

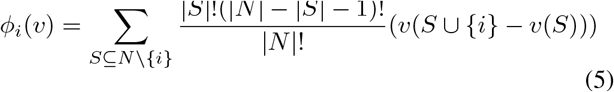

### (5) SHAP formula; info from (Cotra, 2019)

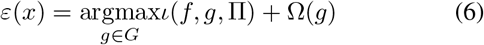

### (6) LIME formula; info from (Sharma, 2018)

When going about getting visualizations, the input image is segmented first. This aids in not having to explain every pixel.

**Fig. 16.**
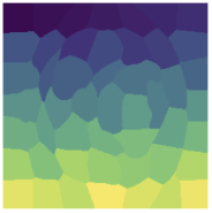
Segmented image (VGG16)

Afterward, the top prediction for the inputted image is received. In this case, it was malignant. Finally, the visualizations are produced.

**Fig. 17.**
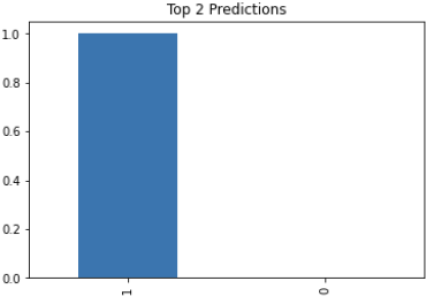
Top prediction for the inputted image (VGG16)

**Fig. 18.**
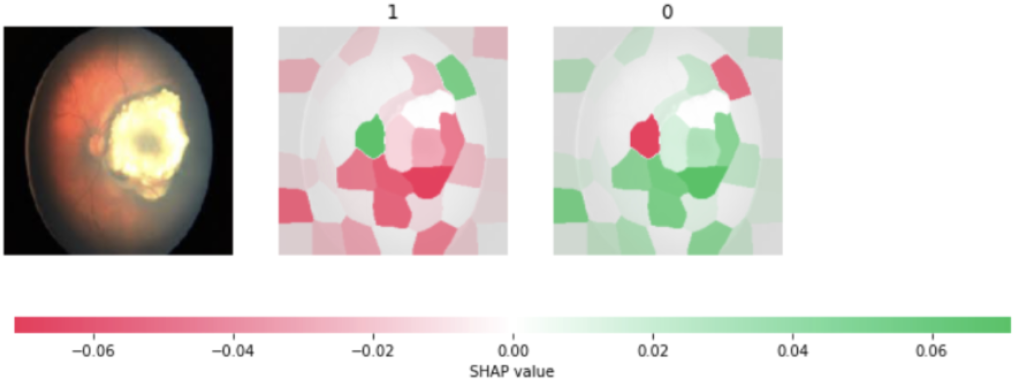
SHAP visualizations for VGG16 (performed during preliminary testing stage)

## IV. Conclusion

In this project, multiple transfer learning architectures were analyzed: VGG16, VGG19, ResNet50, Inception v3, and ResNet50. VGG19 is an extension of VGG16. VGG19 contains 19 layers (16 of which are convolutional layers), and VGG16 contains 16 layers (13 of which are convolutional). With regards to Inceptionv3 and Xception, Xception is an extension of Inception v3 but with depthwise separable convolutions. Inception v4 was considered but was disregarded due to the high-loss results of Inception v3. Accuracy and loss were used as metrics to measure if there was overfitting in models and/or any major flaws, but DsC and IoU were the indicators of the feasibility of each model to be used for RB detection. ResNet50 had the largest concerns for overfitting after VGG16 and VGG19. Concerns of overfitting were mitigated by utilizing separate, unseen testing datasets and early stopping.

**Fig. 19.**
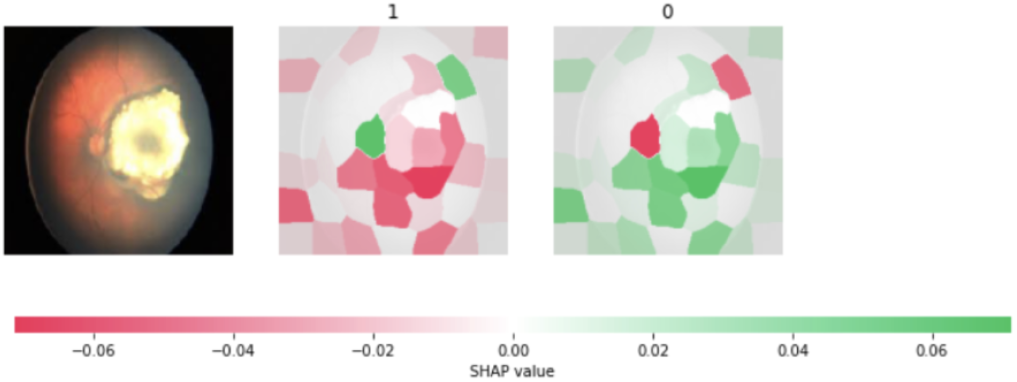
SHAP visualizations for VGG19

**Fig. 20.**
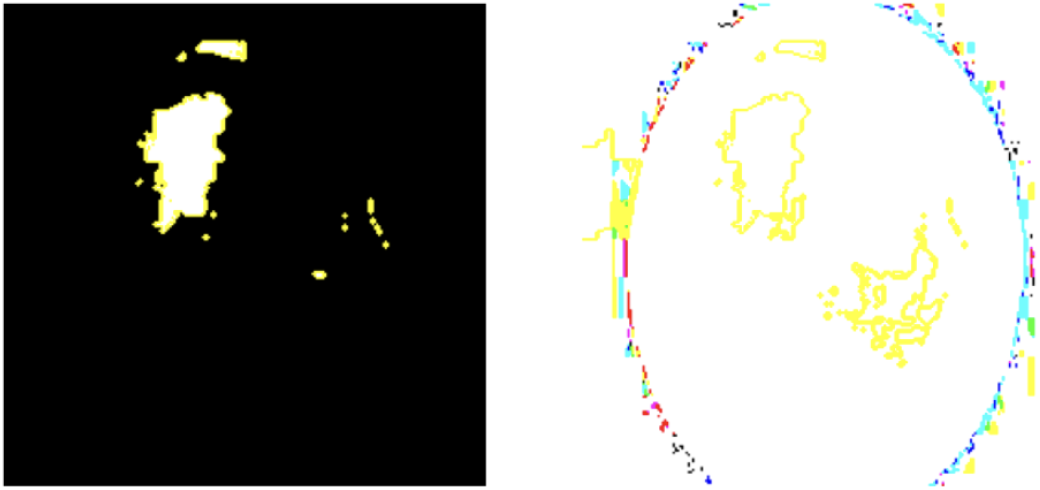
LIME visualizations for VGG19

It was deduced that VGG16 has the best viability in diagnosing RB with a 0.9890 DSC and 0.9787 IoU value. VGG19 was a close runner-up with a 0.9885 DSC and a 0.9786 IoU value. Inception v3 had a 0.9663 DSC and 0.9374 IoU value, with Xception being a close runner-up to that model with DSC and IoU values of 0.9647 and 0.9371, respectively. Finally, ResNet50 performed the worst with a 0.8400 DSC and 0.7052 IoU value.

SHAP was applied to VGG16 and were able to gain better insight into which contours of the image caused the diagnosis that the model gave for a single image. Since VGG19 was a close runner-up, SHAP and LIME were also run on the results of that model for a single image and the model was able to produce interesting visuals making it easier to understand how the model viewed the image that was inputted.

Through the use of metrics such as DSC and IoU, which are specifically designed for medical image classification, and by comparing popular pre-trained image processing architectures, the effectiveness of these models can be evaluated in identifying tumor features. Additionally, by employing XAI techniques such as SHAP and LIME, the transparency of the decision-making process of these models can be enhanced and promote trust in AI among clinicians. These combined efforts can improve the clinical applicability of deep learning models and assist doctors in possibly identifying novel tumor features.

## Data Availability

All malignant image data is available at: https://www.mathworks.com/matlabcentral/fileexchange/99559-retinoblastoma-dataset
Benign images were scraped from multiple publicly available sources and the full benign-malignant dataset is available upon reasonable request to the authors.

## V. Future Research

Artificial intelligence in medicine often faces one common challenge; accurate data to develop effective models. One way to improve the architectures’ accuracy is by acquiring data from previous deep-learning studies on RB. By using this data to feed the models larger datasets, it can be ensured that the models are trained with large, high-quality datasets. To acquire this data, it will be ensured that research ethics guidelines are followed and the necessary licensing is obtained. A correspondence was already made to a previous deep-learning study that utilizes 36,623 images (Zhang et al., 2022). By incorporating the aforementioned data into the models, it is possible to improve the accuracy of the models, resultantly increasing the clinical viability of this AI solution for diagnosing RB in LMICs.

One hanging end of this project is creating a cost-effective device to collect fundus imaging and data for biomarkers, alongside connecting fundus imaging and biomarker data. Fundus imaging is an expensive technique utilized to diagnose ocular diseases and is not very accessible. By finding a way to train the models with iPhone images, which MDEyeCare has explored the feasibility of, this goal can be achieved. In addition, inexpensive methods to collect HIPAA-compliant data are discovered, algorithmic efficiency is maximized to decrease operating costs, and the model is trained with images and possibly biomarkers; it is possible to create a multimodal approach to maximize this solution’s applicability to LMICs. Overall, by incorporating data from existing studies, creating cost-effective data-collecting devices, exploring the use of biomarkers, and following HIPAA compliance guidelines, it is possible to increase the affordability and efficiency of medical models. These efforts can ultimately improve patient outcomes in LMICs and decrease the disparity in access to high-quality medical care.

## Notes

### Competing Interest Statement

This work has been submitted to the IEEE for possible publication. Copyright may be transferred without notice, after which this version may no longer be accessible.

### Funding Statement

This study did not receive any funding.

### Author Declarations

All malignant image data is available at: https://www.mathworks.com/matlabcentral/fileexchange/99559-retinoblastoma-dataset Benign images were scraped from multiple publicly available sources and the full benign-malignant dataset is available upon reasonable request to the authors.

